# Clinical Benefit of Vagus Nerve Stimulation in Intractable Epilepsy: A Systematic Review and Meta-analysis

**DOI:** 10.1101/2025.05.10.25327360

**Authors:** Made Agus Mahendra Inggas, Kennytha Yoesdyanto, Edeline Samudra, Prudence Wirajaya, Nathan Muliawan

## Abstract

**Background:** Intractable or drug-resistant epilepsy (DRE) is a condition where seizures cannot be adequately controlled through antiepileptic medications. In the setting where resective surgery is ineffective, neuromodulation therapy, or vagus nerve stimulation (VNS), is a safe and approved solution. Nonetheless, the efficacy has yet to be clear. We conducted this systematic review and meta-analysis to evaluate the clinical benefit and response of VNS on seizure frequency reduction in intractable epilepsy.

**Methods:** Four databases (PubMed, Elsevier, Google Scholar, Neurology Journals) were searched from inception to November 2024. RCTs and observational studies that analyzed the effect of vagus nerve stimulation in intractable epilepsy patients were included. Review manager (RevMan 5.4) was used for data analysis with random effects model based on heterogeneities.

**Results:** Five cohort studies (three prospective and two retrospective) were included in the quantitative analysis, involving 244 participants with intractable epilepsy. The pooled analysis revealed a significantly increased likelihood of seizure reduction with VNS (RR = 13.55, 95% CI = 4.95-37.05; p <0.001). Adverse events, reported in three studies, were generally mild to moderate. Two studies assessing the relationship between seizure type and VNS response consistently demonstrated a better response in cases of generalized epilepsy. One study found a positive response of VNS therapy after prior surgery in focal resection group (>60%), followed by corpus callosotomy (33%). However, no study reported a significant reduction in AED usage following VNS therapy.

**Conclusion:** VNS is considered a favorable therapy for patients with intractable epilepsy, notably in generalized epilepsy.

## Introduction

Epilepsy is a chronic neurological disorder caused by sudden abnormal synchronous neuronal activity in the brain, leading to temporary brain dysfunction and characterized by recurrent epileptic seizures.^1^ Globally, epilepsy affects 1–5% of the population, approximately 50 million people worldwide, and is common in both children and adults. In children, epilepsy is typically caused by genetic factors, malformations, or perinatal insults, while in adults, it is usually due to neuronal degeneration, infections, trauma, or tumors.^2^ According to WHO reports, around 30–40% of epilepsy patients experience failure to control seizures, known as intractable or refractory epilepsy, which is commonly caused by drug-resistant epilepsy (DRE).^1,3^ The International League Against Epilepsy (ILAE) defines DRE as uncontrollable seizures despite the use of two or more adequately dosed and well-tolerated antiepileptic drugs (AEDs), either as monotherapy or in combination.^4^

Patients with DRE are at high risk of premature death, with a mortality rate five times higher than those with well-controlled seizures. The most common cause of death is sudden unexpected death in epilepsy (SUDEP).^5^ Therefore, DRE patients require non-pharmaceutical approaches, such as surgical intervention, neuromodulation, and a ketogenic diet. Although epileptic focus resection can be curative, not all patients are eligible for resective epilepsy surgery due to factors such as multifocal seizure origins, high risk of functional deficiency due to the proximity of the epileptogenic zone to the eloquent cortex, or the inability to localize the epileptic focus. Even among the 30–50% of patients who undergo epileptic focus resection, seizures may persist due to surgical failure.^1,6^

Neuromodulation is considered an optimal treatment for patients unsuitable for resection surgery or when resection surgery fails to achieve seizure-free status.^7^ There are three neuromodulation options to adjunct AEDs in treating DRE cases: vagus nerve stimulation (VNS), deep brain stimulation (DBS), and responsive stimulation. These neurostimulators deliver regular electrical pulses to the brain to reduce irregular electrical abnormalities that cause seizures.^2,3^ VNS is an adjunct therapy approved by the American Food and Drug Administration (FDA) since 1997 to reduce seizure frequency in patients aged ≥12 years. In 2017, the FDA also approved VNS as a safe and effective treatment for patients aged ≥4 years.^1,8^ VNS has been used to manage neurological conditions such as epilepsy, depression, tinnitus, schizophrenia, and other epilepsy comorbidities.^1^ Prior studies have shown that more than 50% of patients with a VNS stimulator can achieve ≥50% seizure reduction.^7^ Furthermore, earlier studies observed a clear trend of lower mortality rates in DRE patients two years after VNS implantation.^5^

Electrical stimulation of the vagus nerve causes significant changes in cerebral blood flow to areas such as the thalamus, hypothalamus, insula, amygdala, hippocampus, parahippocampus, and cerebellum.^7^ Epilepsy is thought to arise from cerebral hyperemia, and VNS, which can compress the carotid arteries, may reduce cerebral blood flow and modulate cortical activity.^9^

The vagus nerve consists of two asymmetric vagus nerves. The right vagus nerve primarily innervates the sinoatrial (SA) node and carries parasympathetic fibers that innervate the cardiac atria, while the left vagus nerve primarily innervates the atrioventricular (AV) node.^9^ As a result, the left vagus nerve is the typical site for implantation, while right vagus nerve implantation is generally avoided due to the higher risk of sinus bradycardia, asystole, or other cardiac side effects.^2,5^ However, in certain cases where left-sided VNS is not feasible, such as due to neck irradiation or infection, implantation on the right vagus nerve can be performed, with previous studies showing it to be 85% safe.^2,10^

VNS can be divided into classic (invasive/traditional) VNS and transcutaneous (non-invasive) VNS.^11^ Invasive VNS targets both afferent and efferent fibers of the vagus nerve and requires surgical implantation, whereas non-invasive VNS is typically administered through the ear and primarily targets afferent fibers via the auricular branch of the vagus nerve.^12^ However, only invasive VNS has been FDA-approved as an adjunctive therapy for DRE patients, while non-invasive VNS is approved for refractory migraines and cluster headaches.^13^ Invasive VNS has shown promising results in epilepsy cases where resective surgery is not a viable option.^8^

Previous studies regarding the efficacy of VNS in reducing seizure frequency amongst patients with intractable epilepsy have demonstrated somewhat inconsistent results, with some exhibiting significant reductions, while the others displaying insignificant reductions or adverse outcomes.^14–16^ Conflicting results regarding VNS implantation and the requirement of AEDs were also noted. Such gaps between these results led to the necessity to further evaluate the effects and efficacy of VNS towards seizure reduction in intractable epilepsy.

## Methods

### Search and Study Selection

A systematic review was done through PubMed, Elsevier, Google Scholar, and Neurology Journals databases investigating the effect of vagus nerve stimulation in intractable epilepsy patients without publication date limits. The keywords used in various combinations were: (“Vagus nerve stimulation” OR “Vagal nerve stimulation”) AND (“Intractable epilepsy” OR “Drug-resistant epilepsy” OR “Epilepsy surgery”).

The articles were collected using PRISMA diagram and three reviewers screened the titles and abstracts of each study (Figure 1).^17^ Subsequently, the selected studies were reviewed and assessed for eligibility using PICOS (Participants, Intervention, Comparisons, Outcome, Studies) analysis shown in Table 1. We included studies that: i) investigated subjects with intractable epilepsy or presumptive drug-resistant epilepsy (DRE), ii) evaluated interventions using invasive vagus nerve stimulation (VNS), and iii) used randomized controlled trials (RCTs) or comparative observational studies (both prospective and retrospective). We excluded studies that: i) had incomplete reporting or lacked sufficient data on the impact of VNS on seizure control, ii) were not available in English, or iii) had a JADAD score below 3, to ensure the reliability and accuracy of the data, as low-quality evidence may compromise the validity of the meta-analysis results.

**Table 1.**
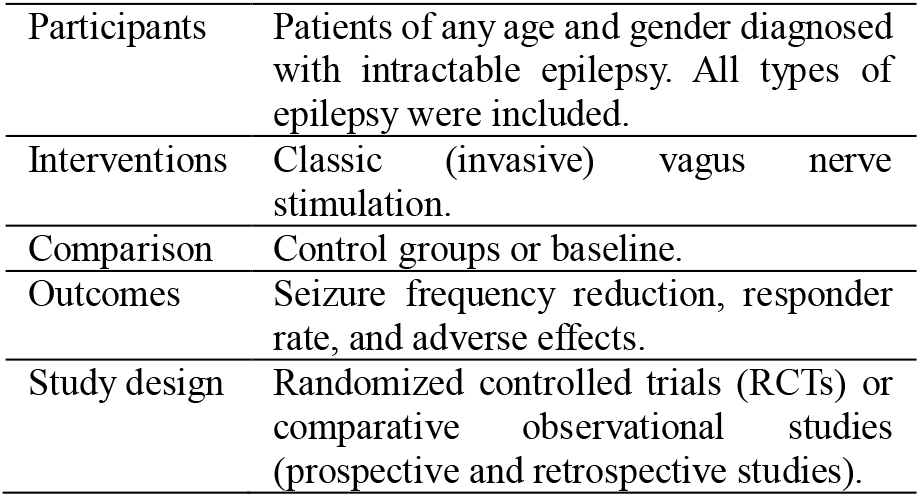
Inclusion criteria in terms of PICOS.

**Figure 1.**
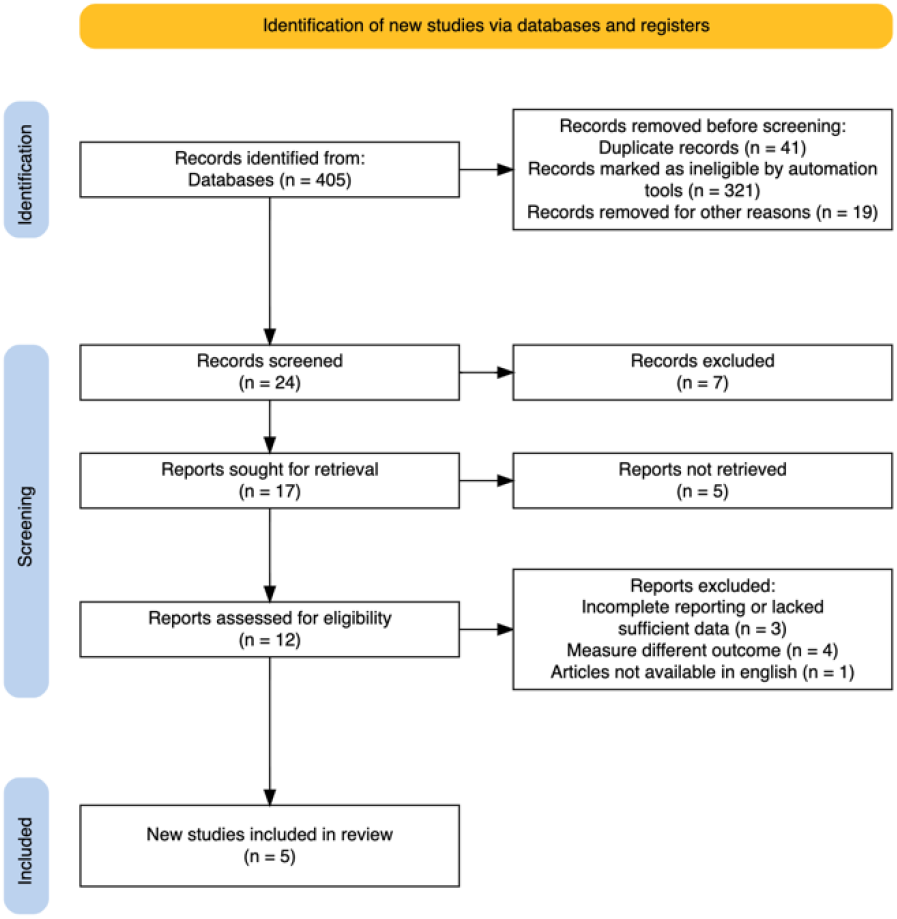
Preferred Reporting Items for Systematic Reviews and Meta-Analyses (PRISMA) flow diagram of study selection

### Type of Intervention

The intervention described in the included studies involved the implantation of vagus nerve stimulation (VNS) devices in conjunction with antiepileptic drugs (AEDs) for patients with intractable epilepsy who were either ineligible for epilepsy surgery or had not achieved seizure control even after surgical resection. The primary outcomes of this study included seizure frequency reduction and responder rate in response to VNS stimulation. Secondary outcomes included the association of epilepsy types and, if applicable, prior surgical resection types with VNS response.

### Data Extraction and Analysis

The risk of bias in each study was assessed using the Risk of Bias in Non-Randomized Studies of Interventions (ROBINS-I) tool.^18^ A random-effects meta-analysis was conducted using Review Manager (RevMan) 5.4.1 software, with results expressed as risk ratios (RRs).

For studies with binary outcomes reporting paired data (e.g., comparing post-intervention results to baseline within the same patient group) that contained zero counts in one or more cells of a 2×2 table, a continuity correction of 0.5 was applied to prevent undefined calculations when calculating RRs. However, since RevMan 5.4.1 does not support decimal values in cell counts, the zero-cell correction was adjusted by multiplying all counts by 2 before applying the correction. Specifically, the correction of 0.5 was converted to 1 and added to the relevant 2×2 table cells.^19^

The included studies were compared and assessed for their level of heterogeneity. Studies with similar methodologies, treatment duration, and outcome measures were grouped together to ensure comparability. Subsequently, we evaluated each study to determine whether their data were comparable, and any significance differences were noted for subgroup analysis, ensuring that the included studies were as similar as possible, thus reducing bias and increasing the reliability of the results. The I^2^ statistic was used to estimate the degree of heterogeneity among the included studies, with an I^2^ value of ≥50% interpreted as indicating statistical heterogeneity.

## Results

### Search Results and Study Characteristic

The literature search yielded 405 articles, of which 5 studies met the inclusion criteria and were included in this meta-analysis. These studies were published between 2008 and 2023, consisted of three prospective cohort studies and two retrospective cohort studies, encompassing a total of 244 patients with intractable epilepsy. The ages of the included patients ranged from 3 to 63 years, with the duration of epilepsy spanning 0.3 to 39 years.

Four studies included patients with both focal and generalized seizure types, while one study did not specify the types of seizures among its participants. Similarly, four studies included patients who had undergone prior epilepsy surgery that failed to achieve seizure control, while one study did not provide information about prior surgeries.

Regarding the intervention, four studies detailed the current levels of VNS provided to patients. Stimulation typically started at 0.25–0.5 mA and was gradually increased to a comfortable and tolerable level, with a mean current exceeding 1.0 mA. One study, however, did not report the VNS intensity.

All studies had a follow-up period of at least 24 months, minimizing time-varying confounders. Seizure frequency reduction and responder rates were assessed as primary outcomes and compared to baseline measures in all studies. Additionally, three studies evaluated adverse effects of VNS, two studies investigated the association of VNS with the number of AEDs required, three analysed the relationship between seizure types and VNS response, and one analysed the association between prior epilepsy surgery type and VNS response. The demographic characteristics of each study are summarized in Table 2.

**Table 2.** Demographic characteristics of each study.

| First author, year | Sample size | Mean age (years) | Study design | Seizure type | Seizure duration (years) | Epilepsy surgery type | VNS current | Follow-up duration | Comparator | Frequency reduction | Responder rates | Side effects & adverse events |
| --- | --- | --- | --- | --- | --- | --- | --- | --- | --- | --- | --- | --- |
| Lee et al., 2008<sup>20</sup> | 11 | 29.82 (ranged 13-50) | Prospective study | Complex partial seizure (72.2%), absence seizure (27.3%) | 4.3 (ranged 0.9 to 11.6) between first epilepsy surgery and VNS | Left temporal lobectomy (40%), corpus callosotomy (40%), corpus callosotomy with topectomy (10%) | 1.08 mA (range: 0.5-1.75 mA) | 24 months | Baseline | 97.20% | 100% (54.5% became seizure-free, 44.5% showed reduction $\geq 50\%$) | Hoarseness (2 patients; 18%) and neck pain (1 patient; 9%) during stimulation |
| Muller et al., 2010<sup>21</sup> | 26 | 19 (median of 3-34 years) | Open-label prospective study | Focal epilepsy (57.7%), lennox-gastaut syndrome (26.9%), spasm seizure (3.8%), progressive myoclonic epilepsy (3.8%), unclassified form of symptomatic generalized epilepsy (7.7%) | Ranged 0.3 to 19 | N/A | 1.5 mA | 24 months | Baseline | 23% at first year, 22% at the second year | 50% at first year and 30% at the second year | Most common: alteration of voice during stimulation (95%), coughing (35%), radiating tingling sensation (30%), and hoarseness (20%) during stimulation<br>Less common: vomiting (12%), insomnia (12%), loss of weight (8%), loss of appetite (4%), and paraesthesias (4%) |
| Vale et al., 2011<sup>22</sup> | 37 | 29.1 (ranged 5-63) | Prospective study | N/A | N/A | Focal resection (31 patients with 6 patients had two different surgical sites: temporal lobectomy (64.86%), frontal lobectomy (32.43%), occipital (2.7%); corpus callosotomy (16.22%) | Start 0.25 mA and increased gradually until comfortable level was reached | >18 months, mean follow-up 61.7 months (5 years) | Baseline | 64.9% achieved reduction <30%<br>24.3% achieved reduction 30-60%<br>10.8% achieved reduction >60% | 10.80% | N/A |
| Shan et al., 2022<sup>23</sup> | 45 | 27.9 (ranged 5-54) | Retrospective study | Focal (37.8%), generalized 62.2%) | 11.8 (ranged 0.25-39) | history of epilepsy surgery (4.4%) | 1.75 mA (start 0.25 mA and increased in increments of 0.25-0.5 mA, with maximum intensity of 2.5 mA) | 5-60 months | Baseline | After 3, 6, 12, 24, 36, 48, 60 months:<br>24.2%, 36%, 42.3%, 45.7%, 46.3%, 58%, 70.3%, respectively;<br>24.4% had 50-79% reduction, 24.4% had 80-100% reduction | 44.4% after 24 months and 48.9% after 60 months; 8.9% became seizure-free | 57.8% experienced adverse events: hoarseness (22.2%), discomfort at the surgical stie (11.1%), coughing (8.9%), nausea (4.4%), paresthesia (4.4%), dyspnea (4.4%), cervical muscle spasm (2.2%) |
| Imura et al., 2023<sup>24</sup> | 125 | 29.2 (± 15.4) | Retrospective study | Focal onset epilepsy (51.64%), focal to bilateral tonic-clonic epilepsy (30.52%), epileptic spasms (10.33%), generalized epilepsy (7.51%) | 16 (± 12.9) | 42% had prior epilepsy surgery | Model of VNS (103/105: 106 = 85: 40) | 24 months | Baseline | At 6 months: 28% achieved reduction of at least 50%.<br>At 12 months: 41% achieved reduction of at least 50%.<br>At 24 months: 52% achieved reduction of at least 50%. | 56% | N/A |

N/A=not applicable, VNS=vagus nerve stimulation

### Risk of Bias and Meta-analysis Results

The risk of bias, assessed using the ROBINS-I tool as shown in Table 3, revealed that three studies demonstrated “low risk” in all areas. One study showed a “moderate risk” in the area of “bias due to confounding”, and another study showed a “moderate risk” in the area of “bias in classification of interventions”.

**Table 3.**
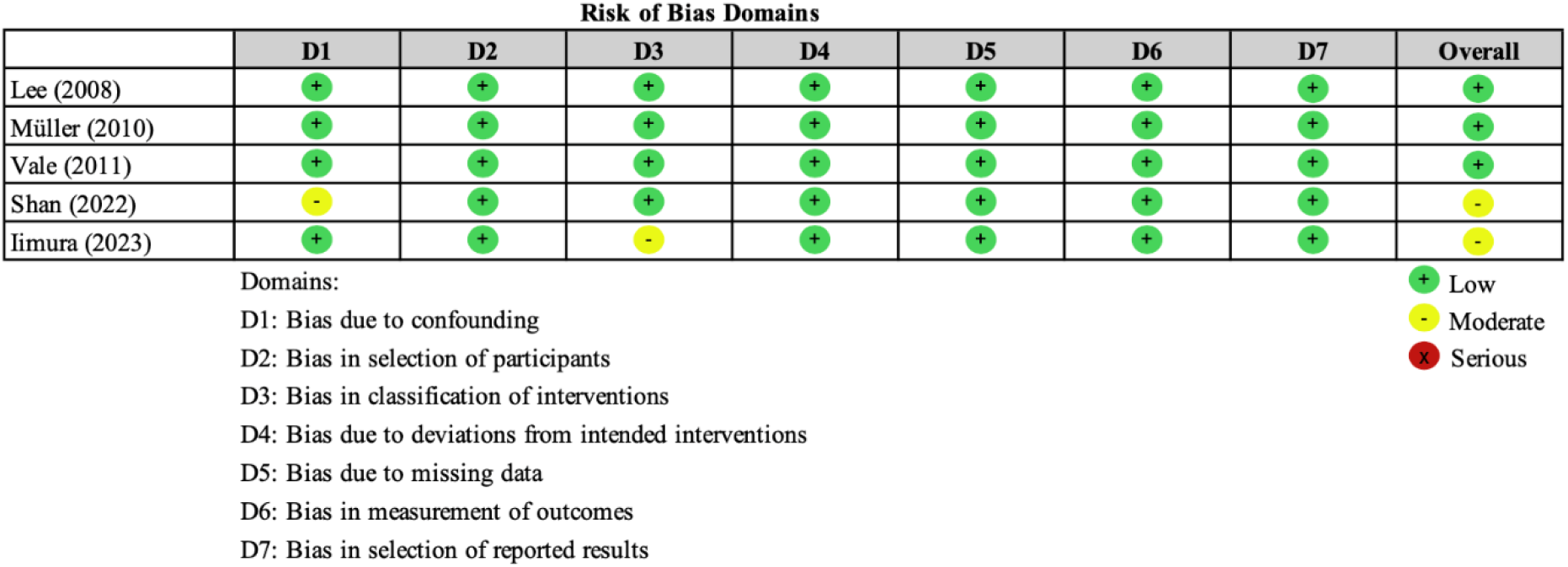
Risk of Bias in Non-Randomized Studies of Interventions (ROBINS-I)

| Risk of Bias Domains |  |  |  |  |  |  |  |  |
| --- | --- | --- | --- | --- | --- | --- | --- | --- |
|  | D1 | D2 | D3 | D4 | D5 | D6 | D7 | Overall |
| Lee (2008) | + | + | + | + | + | + | + | + |
| Müller (2010) | + | + | + | + | + | + | + | + |
| Vale (2011) | + | + | + | + | + | + | + | + |
| Shan (2022) | - | + | + | + | + | + | + | - |
| Iimura (2023) | + | + | - | + | + | + | + | - |

Domains:
D1: Bias due to confounding
D2: Bias in selection of participants
D3: Bias in classification of interventions
D4: Bias due to deviations from intended interventions
D5: Bias due to missing data
D6: Bias in measurement of outcomes
D7: Bias in selection of reported results
+ Low - Moderate × Serious

The I^2^ statistic from this study showed that Tau^2^ = 0.39 indicates moderate variability in the true effect sizes across studies, while I^2^ = 30% indicates low heterogeneity. This suggests that most of the variation is likely attributable to chance, implying relatively consistent effects across studies. Additionally, Chi^2^ = 5.70, df = 4 (P = 0.22) indicates that the differences between study results were not statistically significant, and the observed variability is more likely due to random sampling error rather than actual differences in study outcomes, making the studies reasonably comparable. However, the test may lack statistical power due to the small number of studies included.

A wide confidence interval (CI) could be due to small sample sizes or variability in individual study outcomes. However, the pooled proportion, represented by the CI range of 4.95 to 37.5, suggests that VNS stimulation is likely to have a positive effect.

### Primary Outcome

The main outcomes to determine the efficacy of VNS in intractable epilepsy were seizure frequency reduction and responder rate (Figure 2). The result of this meta-analysis showed a pooled ≥50% seizure reduction rate of 46.9%, with RR of 13.55, indicating intervention with VNS significantly increased the likelihood of achieving the outcome.

**Figure 2.**
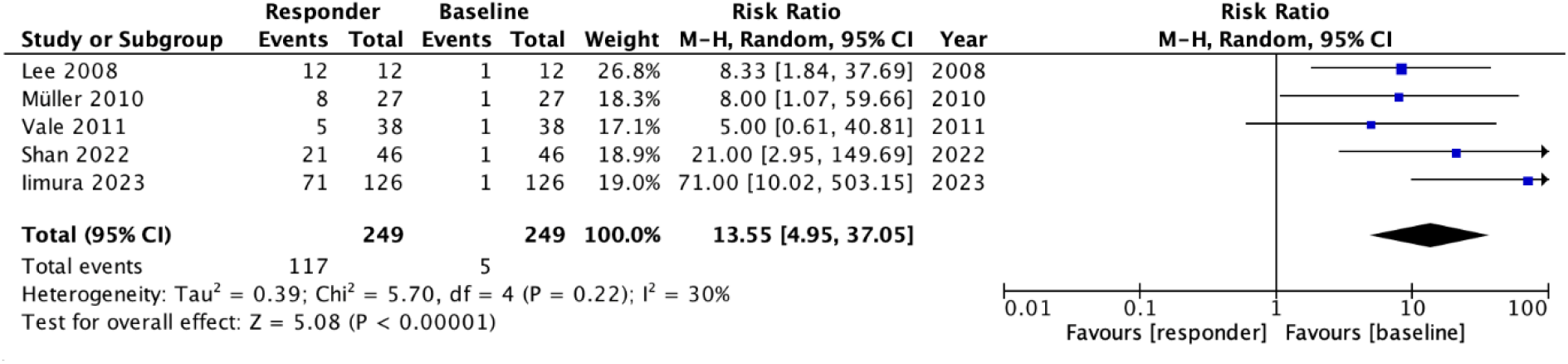
Forest plot. Responder rate of VNS stimulation in intractable epilepsy compared to baseline. Events represent the number of patients who achieved ≥50% seizure reduction.

Two studies reported responder rates, defined as achieving ≥50% seizure reduction, exceeding 50%. Specifically, Lee et al. observed a responder rate of 100%, with 54.4% of patients becoming seizure-free and a mean seizure frequency reduction of 97.2%. Iimura et al., which included the largest number of participants, reported a responder rate of 56% after 24 months but did not specify detailed seizure frequency reductions. Their data showed that 28% and 41% of patients achieved at least a 50% reduction at 6 and 12 months, respectively.

Two other studies reported responder rates of ≥30%, with Müller et al. reporting a 30% responder rate and Shan et al. reporting 44.4%. Müller et al. recorded a mean seizure frequency reduction of 22% in the second year, while Shan et al. found that 24.4% of patients experienced a 50–79% reduction, and another 24.4% achieved an 80–100% reduction. In contrast, Vale et al. reported only 10.8% of patients achieving >60% reduction but they used a different cut-off, defining reduction ranges of 30–60% in 24.3% of patients and <30% in the remaining 64.9%.

### Secondary Outcome

Adverse events were assessed in three studies and were generally mild to moderate. Commonly reported adverse effects during activation of VNS included hoarseness, coughing, and discomfort, with no reports of permanent complications. One study also noted additional events such as discomfort at the surgical site, paraesthesia, dyspnea, and cervical muscle spasm.

Two studies evaluated the association of VNS with the number of antiepileptic drugs (AEDs) required. They did not find VNS significantly reduced AED use. Müller et al. reported that only 4% of patients could taper one or more drugs, while Vale et al. noted an increase in patients requiring only one AED, from 2.7% pre-VNS to 13.5% post-VNS. However, there was also an increase in patients requiring four or more AEDs, from 16.7% pre-VNS to 29.7% post-VNS.

Out of three studies that investigated the relationship between seizure types and VNS response, two studies (Iimura et al. and Müller et al.) reported better responses in cases of generalized epilepsy, while Shan et al. did not find significance differences when comparing non-focal to focal seizures. Iimura et al. identified a statistically significant association (p=0.03) and suggested that the involvement of the thalamus in seizure onset might be the primary mechanism of VNS efficacy in generalized seizures. However, they did not specify which type of generalized seizure benefited most, likely due to small the number of patients. Müller et al. observed a statistically significant response in the generalized tonic–clonic seizure (GTCS) group (p=0.04) compared to all other seizure types. Nonetheless, while there was better seizure reduction in other non-focal epilepsy cases, these findings were not statistically significant. Müller et al. also observed a moderate response across syndromes, with 15% improvement on the Clinician’s Global Impression (CGI) scale at the second year in patients with Lennox-Gastaut syndrome (LGS), reflecting a reduction in seizure frequency.

Vale et al. assessed the relationship between prior epilepsy surgery and VNS response. They found that seizure reductions >60% occurred only in patients who had undergone prior focal resection. Among these, 48% reported an improved quality of life, followed by 33% in the corpus callosotomy group reporting similar improvements.

## Discussion

Intractable epilepsy is associated with increased mortality, morbidity, and a decreased quality of life, necessitating treatments beyond AEDs. When surgical resection is not feasible or not effective in achieving seizure reduction, VNS has been approved as a safe and effective treatment for intractable epilepsy cases.^11^

### The Role of VNS in Reducing Seizure Frequency

From this meta-analysis results, the pooled ≥50% seizure reduction rate of 46.9% and RR of 13.55 indicated significant impact of VNS in reducing seizure frequency ≥50% among intractable epilepsy patients.^20–24^ In line with our findings, a meta-analysis focusing on the impact of VNS in the genetic etiology of drug-resistant epilepsy reported a ≥50% seizure reduction rate of 41%.^25^ The results of this meta-analysis are based on a two-year follow-up period and align with findings from a large review involving 362 Japanese patients. That review reported that the 50% responder rate improved from 55.8% in the first year to 57.7% after two years, and slightly declined to 53.8% after three years.^26^ In contrast to our findings, a retrospective matched pairs case-control study comparing the long-term outcomes between patients receiving best available drug therapy (BDT) with VNS to those receiving BDT without VNS failed to demonstrate the efficacy of VNS in reducing seizure frequency. This study identified a significant increase in the frequency of multifocal epilepsies (p=0,02) and complex partial seizures (p=0,046), along with an insignificant trend towards increased total seizures (p=0,07) among patients receiving BDT with VNS.^15^

The primary neurobiological mechanism behind the antiepileptic effects of VNS remains incompletely understood. However, several potential mechanisms have been proposed, including structural effects on the brainstem, forebrain, and limbic system via the nucleus of the tractus solitarius (NTS), as well as functional effects on neurotransmitters, neuroelectrophysiology, and inflammation in the central nervous system.^1,6^

VNS can increase concentrations of neurotransmitters that suppress seizures, such as norepinephrine, serotonin, and gamma-aminobutyric acid (GABA), while reducing levels of seizure-triggering neurotransmitters like aspartate.^27^ The vagus nerve, or the tenth cranial nerve, is the longest cranial nerve, consisting of 80% afferent fibers and 10% efferent fibers.^9^ The afferent fibers modulate activity in subcortical and cortical circuitry, with 80% of these fibers projecting to the NTS. The NTS then sends monosynaptic projections to various brainstem regions, such as the parabrachial nuclei (PBN), noradrenergic locus coeruleus (LC), serotonergic raphe nucleus (RN), periaqueductal gray matter, and cerebellum. The LC is crucial for generating noradrenergic innervation, and transient VNS lasting 0.5 seconds can trigger rapid, periodic neuronal activity in the LC, promoting the release of norepinephrine. VNS also stimulates the RN, enhancing serotonergic cortical innervation, while the PBN connects to the insula, thalamus, hippocampus, amygdala, and limbic system.^1,9^ Prior study also reported that VNS increases levels of inhibitory GABA in the cerebrospinal fluid and reduces excitatory glutamate signaling, thereby lowering susceptibility to chemically induced limbic motor epilepsy.^1^

VNS also regulates central nervous system electrophysiology and reorganizes the pathologically hypersynchronous brain networks of DRE patients.^28^ VNS modifies neuronal activity in the amygdala and hippocampus by increasing the protein content at the postsynaptic density (PSD). The PSD is a functional protein complex located on the postsynaptic membrane, playing a crucial role in the structural, functional, and plasticity processes of excitatory synapses in the central nervous system. Increased VNS intensity can further elevate the firing rate in the amygdala.^1^ A study by Li et al. found that VNS stimulation significantly reduced functional connectivity in alpha and theta bands, increased network efficiency, and improved cognitive function in DRE patients.^28^ Thus, VNS modifies and alters the composition of excitatory synapses, inducing desynchronization of cortical electrical activity in epileptic patients.

VNS also exhibits anti-inflammatory actions, regulating inflammatory factors in the central nervous system and inflammation-related diseases such as sepsis, arthritis, and excessive ischemia-reperfusion injury. Since epilepsy can be triggered by inflammation and immune system activation, VNS, which reduces proinflammatory cytokines and tryptophan metabolites, plays a role in mitigating epileptic seizures.^1,9^

Lastly, VNS induces significant changes in regional cerebral blood flow in DRE patients, particularly in the thalamus and cerebral cortex.^28^ Functional imaging studies have shown that VNS activates and increases cerebral blood flow in the bilateral thalami, hypothalami, insular cortices, and inferior cerebellar hemispheres.^9^

### Intensity of VNS

Four studies included in this meta-analysis reported a mean current exceeding 1.0 mA, typically initiated at 0.25–0.5 mA and gradually increased to a comfortable and tolerable level.^20–23^ This is consistent with findings from a review by Cramer et al., which also reported higher seizure frequency reductions (22-43%) with high-frequency stimulation (>20 Hz) and an average current of 1.3 mA, compared to low-frequency stimulation (1 Hz) or baseline seizure frequency.^29^ Roosevelt et al. (2006) found that VNS at 1 mA increased the extracellular concentration of norepinephrine in the bilateral cortex and hippocampus. In contrast, stimulation at 0.5 mA significantly increased norepinephrine concentration only in the hippocampus, with no increase observed at stimulation levels below 0.5 mA.^30^ VNS is commonly administered at levels of 1–2 mA, with a frequency of 20–30 Hz, a pulse width of 150–500 µs, and an on-time of 30 seconds followed by an off-time of 3–5 minutes.^7^ While the VNS device can produce stimulation up to mA, excessive output may decrease its efficacy and cause laryngeal complications such as hoarseness, dysphagia, and coughing.^7,9,31^

### Impact of VNS on the Number of AEDs Required

Two studies included in this meta-analysis did not find a significant impact of VNS on reducing the number of AEDs.^21,22^ Consistent to our findings, a retrospective study investigating the therapeutic effects of VNS among children with drug-resistant epilepsy reported unremarkable or minimal impact on the reduction of AEDs. This study involved a two-year follow-up, reported that 13.6% of patients had a reduced AED regimen, from requiring three or more AED types to only one or two. However, 5% of patients had an increase in their AED regimen instead, from requiring only one or two types, to requiring three or more. No change in AED regimen was observed in 75% of patients. This suggests the absence of statistical significance in the change of AED regimens following VNS implantation.^16^

In contrast to our findings, a study reviewing various outcome measures following VNS implantation reported notable reduction in the number of AEDs used. Patients involved in this study were initially treated on average with 5.2 types of AEDs prior to VNS implantation. Following VNS implantation, these patients were reported to be treated on average with 4.2 types of AEDs.^14^ The previously mentioned retrospective matched pairs case-control study yielded similar results. In that study, a larger proportion of patients receiving BDT with VNS required more than two types of AED. It was reported that 10/20 patients receiving BDT with VNS required more than two types of AEDs, while only 2/20 patients receiving BDT without VNS required more than two types of AEDs.^15^

Responses to VNS may vary among each patient, in which some may experience marked reductions in seizure frequency, whereas others show little to no improvement. Such variability suggests that although beneficial in certain patients, VNS may not serve as a replacement for AEDs. VNS often serve as an adjunct to AEDs, to achieve optimal seizure control. In cases where VNS do not sufficiently reduce the use of AEDs, patients typically require adjustments to their medication regimen, potentially involving higher doses which could increase the risk of side effects. Such combined therapy necessitates close and frequent evaluation, to identify interactions between the two modalities and create adjustments to optimize seizure control. If VNS proves ineffective in reducing seizure frequency or the use of AEDs, alternative treatments including surgical resection or deep brain stimulation (DBS) may be considered.^32^

### Association of Seizure Types with VNS Response

Three studies included in this meta-analysis investigated the relationship between seizure types and VNS response in two studies reported better responses in cases of generalized epilepsy, with one of them specifically reported significant response in the generalized tonic–clonic seizure group.^21,23,24^ The response to VNS therapy varies across different seizure types. Major seizures such as focal-to-bilateral tonic-clonic and generalized motor seizures showed better improvement with VNS therapy, with a significant reduction in both seizure intensity and frequency. In contrast, minor seizures such as focal-aware and non-motor/absence seizures exhibited less pronounced responses.^33,34^ Urian et al. reported that 82% of patients with major seizures achieved a favorable response, defined as a ≥50% reduction in seizure frequency after 12 months of VNS therapy, while only 25% of patients with minor seizures achieved a similar response.^34^ Similarly, Moshref et al. found that patients with non-lesional drug-resistant epilepsy (DRE) showed higher rates of seizure reduction and freedom compared to lesional cases. However, lesional groups demonstrated better seizure reduction rates if they had undergone prior epilepsy surgery.^35^ Additionally, conditions such as tuberous sclerosis and posttraumatic epilepsy respond well to VNS.^36^

In contrast to our findings, a review of VNS therapy for focal seizure patients conducted by Panebianco et al. reported an RR for seizure frequency reduction ≥50% of 1.73 (95% CI: 1.13–2.64), indicating a positive impact of VNS on focal seizures. Similarly, Wang et al. observed better outcomes in cases with unilateral interictal epileptiform discharges, while Arcos et al. noted that temporal lobe discharges detected through video-EEG were associated with an early favorable response to VNS therapy.^36,37^

In addition to seizure types, prior studies consistently found that a shorter duration of epilepsy before VNS implantation was strongly correlated with better results. However, neither the age at seizure onset nor the age at VNS implantation showed a significant relationship with outcomes.^36^

### Association of Prior Surgical Resection with VNS Response

One study included in our meta-analysis, conducted by Vale et al., reported that seizure reductions greater than 60% occurred only in patients who had undergone prior focal resection, with 48% of these patients experiencing an improved quality of life.^22^ In contrast to our findings, Wang et al. found no significant association between a history of prior epilepsy surgery and the response to VNS therapy.^36^ Additionally, Vale et al. reported that 33% of patients in the corpus callosotomy group experienced improved quality of life. This aligns with other previous studies reporting better VNS responses in patients with a history of corpus callosotomy.^36^

### Safety Profile

This meta-analysis found that VNS is a safe treatment with a low incidence of adverse events, which are generally mild to moderate and can be resolved. Common adverse effects reported include hoarseness, coughing, and discomfort during stimulation, which were well-tolerated and manageable through adjustments to the output currents. In line with our findings, previous studies indicate that VNS implantation is relatively safe, with a complication incidence of around 2%. Complications may include infection, vocal cord paralysis, postoperative hematoma, or equipment-related issues such as battery replacement, lead fracture, malfunction, and the need for repeat surgery.^1^

### VNS in Comparison with Repeat Epilepsy Surgery

In DRE cases where resective surgery is possible, seizure freedom is achieved in 40–80% of patients. However, one-third of these patients experience recurring or persistent seizures.^38,39^ Postsurgical outcomes depend on the location and extent of the epileptogenic zone, with unfavorable results often attributed to incomplete resection. Persistent seizures may arise from inaccurate localization of the epileptogenic zone due to insufficient presurgical identification, limited resectability due to the involvement of functional areas, or bilateral or multifocal ictal discharge patterns. Recurring seizures may also result from the progression or recurrence of underlying causes such as tumors or the emergence of new epileptogenic regions, as identified in 26% of patients in Kunz et al.’s study.^38,40^ Some patients with genetic epileptogenic substrates may develop new foci even after complete resection due to gene expression.^40^ In such cases, options include adjusting medication regimens, palliative treatments like VNS, or repeat surgery.

The success rates for repeat surgery vary. Kunz et al. found that 71% of patients with refractory focal epilepsy achieved seizure freedom after a second epilepsy surgery.^40^ Similarly, Hu et al. reported that 53.5% of patients achieved seizure freedom, with 83.3% of those with focal or unilateral onset seizures becoming seizure-free, while 78.6% of those with bilateral or multifocal seizures continued to have seizures.^38^ A meta-analysis by Krucoff et al. showed that only 47% of patients undergoing repeat resective surgery achieved seizure freedom.^41^ These findings consistently indicate favorable outcomes for repeated epilepsy surgeries in focal epilepsy, especially temporal lobe epilepsy. However, patients with bilateral seizure onset are more likely to experience postoperative seizure recurrence.^38^ Furthermore, neocortical temporal lobe epilepsy outcomes are less favorable than those for mesial temporal lobe epilepsy due to the difficulty in localizing neocortical epileptic foci.^42^ Abnormal MRI findings can help predict better outcomes by guiding resection margins.^43^

Repeat epilepsy surgery carries risks of complications. Fukuda et al. reported that only one patient experienced complications after the first surgery, compared to 56% of patients (10 out of 18) who experienced complications after additional surgeries.^44^ Hu et al. found that repeat epilepsy surgery led to permanent neurological deficits in 30.2% of patients.^38^ Given the risks, nondestructive therapies like neurostimulation, including VNS, are viable options for patients with epilepsy arising from the neocortex or eloquent cortex.^42^

### Invasive VNS in Comparison with Non-invasive VNS

Transcutaneous VNS (nVNS) has several advantages, such as being noninvasive, less expensive, and causing fewer side effects. However, nVNS only stimulates sensory fibers, whereas classic VNS stimulates both sensory and motor fibers of the vagus nerve.^31^ A summary of the comparison is shown in Table 4.

**Table 4.** Comparison of invasive VNS (iVNS) and transcutaneous VNS (nVNS)^1,9,31^.

|  | iVNS | nVNS |
| --- | --- | --- |
| Point of entry for electrodes | Carotid sheath | Auricle |
| Stimulated division of vagus nerve | Cervical vagus nerve | Auricular branch of vagus nerve |
| Stimulated fibers | Motor and sensory | Sensory |
| Structures activated | Brainstem, thalamus, | Limbic system (mainly |
|  | hypothalamus,<br>insular cortices,<br>cerebellum | posterior<br>cingulate gyrus<br>and<br>hippocampus) |
| Cost-effective | More expensive | Less expensive |
| Side effect (both<br>tolerable and<br>resolvable) | Mild to moderate<br>($\leq 60\%$) | Mild to<br>moderate; less<br>common ($\leq 10\%$) |
| FDA approval | Approved | Not yet |

### Strengths and Limitation

Compared to prior meta-analyses, this study offers several novel contributions that expand the understanding of VNS therapy based on existing evidence base. First, this study emphasizes the clinical benefit of VNS across various types of epilepsy, providing insights into its broader therapeutic impacts. Second, with no significant association found between age and VNS response, combined with the FDA’s approval of VNS for patients aged ≥4 years, this study adopted a broad age inclusion criterion to enhances insight. Third, we evaluated secondary outcomes that may guide personalized treatment strategies for refractory epilepsy, such as the impact of VNS on the number of AEDs required, the association between seizure types and prior surgical resection with VNS response, and adverse effects. Additionally, this study included data on the efficacy of VNS when compared with repeat epilepsy surgery in patients who had previously undergone surgical resection. We also compared iVNS with nVNS, highlighting the latter as a simpler alternative that does not require surgical implantation, though it has not yet received FDA approval.

Some limitations in this study are: First, the limited availability of studies, which resulted in few comparable studies, leading to a small number of included studies and introducing the possibility of sampling error and publication bias. Second, the varied duration of epilepsy among participants may have impacted the current results. Third, it was a pity that comparable RCTs could not be identified and included in this study. Lastly, not all studies provided details regarding VNS current, adverse events, seizure types, or prior epilepsy surgery types. It was also unfortunate that this study could not compare VNS to repeat surgery, as limited studies have addressed this. Further randomized controlled trials are needed to compare VNS with other treatment options, such as surgical resection.

## Conclusion

Intractable epilepsy is associated with an increased mortality rate, suggesting the need for non-pharmaceutical approaches. Vagus nerve stimulation (VNS) has been approved by the FDA, and this study confirmed the efficacy and safety of VNS in intractable epilepsy, with a pooled ≥50% seizure reduction rate of 46.9%, and an RR of 13.55, indicating a significant impact. VNS was associated with a low incidence of adverse effects, which were generally mild to moderate in nature, and typically resolved with minimal intervention. This study also suggests that VNS was particularly effective in patients with generalized motor seizures, which were associated with better treatment outcomes and more significant seizure reduction. Conversely, patients with focal-aware seizures demonstrated less significant responses to VNS and may be better managed through surgical resection. However, a history of previous surgical resection in patients with focal-aware seizures demonstrated improved responses to VNS. The study further recommends VNS for patients who have yet to achieve seizure control despite previous surgical resection, as repeat epilepsy surgery carries a number of potential complications. Therefore, neuromodulation such as VNS with proven efficacy can be considered for drug-resistant epilepsy (DRE) cases.

## Data Availability

All data produced in the present work are contained in the manuscript

## Ethics statements

### Patient consent for publication

Not applicable.

### Ethics approval

Not applicable.

